## Supplemental Materials for "An Explainable Artificial Intelligence Approach for Predicting Cardiovascular Outcomes using Electronic Health Records"

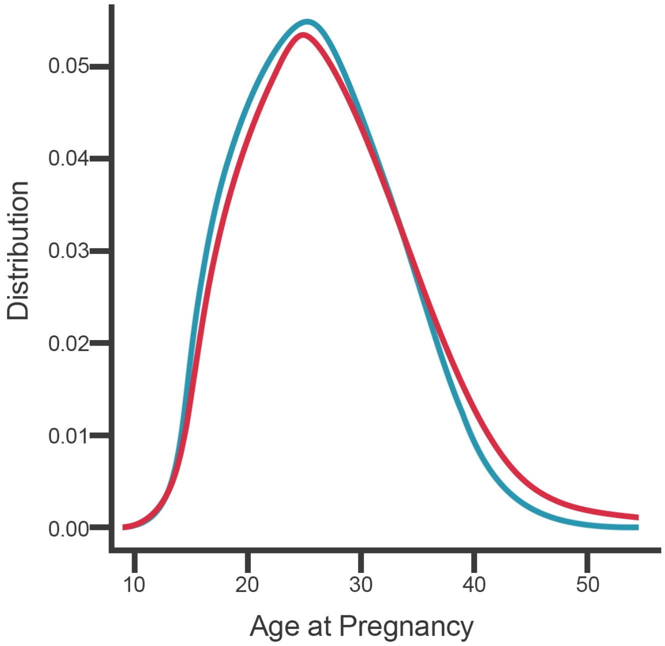


**Supplemental Figure 1. Distribution density plot of mother's age at pregnancy, with and without hypertension complicating pregnancy.** Blue line: mothers with diagnosis of hypertension complicating pregnancy (N=11,523 mothers). Red line: mothers without diagnosis of hypertension complicating pregnancy (N=113,491 mothers).

**Supplemental Table 1. Overview of Utah Data Resource.**

| | |
|---|---|
| Patients | 1,659,372 |
| Patient Encounters | 21,621,986 |
| Billing Codes Associated with Encounters | 165,976,788 |
| Distinct Diagnoses | 12,782 |
| Distinct Procedures | 22,088 |
| Distinct Medications | 2,384 |
| Number of Pediatric Echos | 120,792 |

**Supplemental Table 2. Demographic variables and the Utah Data Resource.**

| GENDER | | ETHNICITY | |
|---|---|---|---|
| Female | 763406 | Not Hispanic | 776547 |
| Male | 712740 | Hispanic or Latino | 152647 |
| Unknown | 116 | Unknown | 547068 |
| ANCESTRY | | | |
| Caucasian | 843275 | Asian | 26144 |
| Unknown/Other | 707593 | African American | 21512 |
| Native Hawaiian or Pacific Islander | 12288 | American Indian or Alaska Native | 8456 |
| FINANCIAL CLASS | | | |
| Commercial | 823504 | Medicaid/Self Pay | 274058 |
| Medicare | 144443 | Other/Unknown | 234257 |

**Supplemental Table 3. Multimorbidity Landscape of Sinoatrial Node Dysfunction (SND) in adults.** Risk and fold-change risk estimates calculated from the multimorbidity network in **Figure 4B** main text. For detailed description of the clinical variables, please refer to supplemental table 5.

| CLINICAL VARIABLES | RISK | FOLD CHANGE |
|---|---|---|
| Male | 0.0047 +/- 0.0001 | 1.01 +/- 0.00 |
| Hispanic | 0.0018 +/- 0.0002 | 0.39 +/- 0.03 |
| Not Hispanic | 0.0048 +/- 0.0001 | 1.04 +/- 0.01 |
| Caucasian | 0.0047 +/- 0.0001 | 1.01 +/- 0.01 |
| African American | 0.0027 +/- 0.0003 | 0.57 +/- 0.07 |
| Medicaid/Self Pay | 0.0018 +/- 0.0001 | 0.40 +/- 0.02 |
| Commercial | 0.0027 +/- 0.0001 | 0.57 +/- 0.01 |
| Medicare | 0.0197 +/- 0.0004 | 4.24 +/- 0.07 |
| Atrial Fibrillation, Male | 0.145 +/- 0.005 | 31 +/- 1 |
| Atrial Fibrillation, Female | 0.145 +/- 0.005 | 31 +/- 1 |
| Atrial Fibrillation, Not Hispanic | 0.143 +/- 0.005 | 31 +/- 1 |
| Atrial Fibrillation, Medicaid/Self Pay | 0.071 +/- 0.010 | 15 +/- 2 |
| Atrial Fibrillation, Commercial | 0.072 +/- 0.004 | 15 +/- 1 |
| Atrial Fibrillation, Medicare | 0.107 +/- 0.004 | 23 +/- 1 |

**Supplemental Table 4. Risks of Cardiac or Nervous System Congenital Anomalies as a Function of Comorbid Clinical Variables.** Risk of cardiac (**Panel A**) or nervous system (**Panel B**) congenital anomalies given the presence of specific clinical variables. Baseline risk and fold change risk calculated using the multimorbidity network in Figure 5 of the main text. For example, a child with a known diagnosis of Down Syndrome has a 25.9-fold increased risk of a cardiac congenital anomaly over the marginal risk of cardiac anomaly. HTN-PREG, hypertension complicating pregnancy (AKA pregnancy-induced hypertension). For detailed description of the clinical variables please refer to supplemental table 5.

**A**

| CLINICAL VARIABLES | RISK | FOLD CHANGE |
|---|---|---|
| Downs | 0.72 +/- 0.02 | 25.9 +/- 0.08 |
| Diaphragm | 0.43 +/- 0.02 | 15.7 +/- 0.8 |
| Nervous | 0.25 +/- 0.03 | 9.2 +/- 0.9 |
| Digestive | 0.18 +/- 0.02 | 6.5 +/- 0.7 |
| Cleft Lip | 0.18 +/- 0.04 | 6.4 +/- 1.5 |
| Eye | 0.12 +/- 0.01 | 4.3 +/- 0.4 |
| Skeletal | 0.11 +/- 0.01 | 3.9 +/- 0.3 |
| Genito-Urinary | 0.09 +/- 0.00 | 3.4 +/- 0.2 |
| HTN-PREG | 0.05 +/- 0.00 | 1.8 +/- 0.0 |
| Skin | 0.04 +/- 0.00 | 1.4 +/- 0.1 |

**B**

| CLINICAL VARIABLES | RISK | FOLD CHANGE |
|---|---|---|
| Cleft Lip | 0.221 +/- 0.037 | 18.4 +/- 3.4 |
| Diaphragm | 0.161 +/- 0.068 | 13.6 +/- 5.9 |
| Downs | 0.124 +/- 0.021 | 10.3 +/- 1.6 |
| Eye | 0.122 +/- 0.015 | 10.1 +/- 1.1 |
| Cardiac | 0.115 +/- 0.010 | 9.6 +/- 0.7 |
| Digestive | 0.110 +/- 0.014 | 9.2 +/- 1.1 |
| Skeletal | 0.102 +/- 0.011 | 8.5 +/- 0.8 |
| Genito-Urinary | 0.061 +/- 0.006 | 5.1 +/- 0.4 |
| Skin | 0.020 +/- 0.003 | 1.7 +/- 0.2 |
| HTN-PREG | 0.016 +/- 0.002 | 1.4 +/- 0.1 |

### Supplemental Table 5. Reference table for EHR coding.

| TERM NAME USED | DESCRIPTION | MATCHED CODES |
|---|---|---|
| Acute CD | Acute Cerebrovascular Disease | CCS DX 7.3.1 |
| Acute Kidney Failure | Acute Kidney Failure | ICD10 N17 |
| Amyloidosis | Amyloidosis | ICD10 E85 |
| AS | Atrial Valve Stenosis | CCS 14.1.7, Echo Derived |
| ASD | Atrial Septal Defect | CCS 14.1.4, Echo Derived |
| Atrial Fibrillation | Atrial Fibrillation | CCS 7.2.9.3 |
| Atrial Flutter | Atrial Flutter | CCS 7.2.9.4 |
| AVSD | Atrio-Venctricular Septal Defect | Echo Derived |
| AVSD unbalanced | Atrio-Venctricular Septal Defect unbalanced | Echo Derived |
| BAV | Bicuspid Aortic Valve | Echo Derived |
| Cardiac | Congenital Cardiac/Heart Defect | CCS 14.1.1, CCS 14.1.10, CCS 14.1.11, CCS 14.1.12, CCS 14.1.13, CCS 14.1.14, CCS 14.1.15, CCS 14.1.16, CCS 14.1.2, CCS 14.1.3, CCS 14.1.4, CCS 14.1.5, CCS 14.1.6, CCS 14.1.7, CCS 14.1.9 |
| Cardiogenic Shock | Cardiogenic Shock | ICD10 R57. |
| Cardiomyopathy | Cardiomyopathy | CCS 7.2.2.1, Echo Derived |
| CleftLip | Cleft Lip | CCS 14.5.2 |
| Coarctation | Coarctation of the Aorta | CCS 14.1.9, Echo Derived |
| Congestive Heart Failure | Congestive Heart Failure | ICD10 I50.4,ICD10 I50.3,ICD10 I50.2 |
| DCM | Dilated Cardiomyopathy | Echo Derived |
| Diaphragm | Congenital Abnormalities of the Diaphragm | CCS 14.5.20 |
| Digestive | Congenital Abnormalities of the Digestive System | CCS 14.2 |
| Downs | Trisomy 21 | CCS 14.5.25 |
| dTGA | d-Transposition of the Great Arteries | Echo Derived |
| Eye | Congenital Abnormalities of the Eye | CCS 14.5.3 |
| Fontan | Fontan Surgery | Echo Derived |
| Genito-Urinary | Congenital Abnormalities of the Genito-Urinary System | CCS 14.3 |
| Glenn | Glenn Surgery | Echo Derived |
| Heart Failure | Heart Failure | ICD10 I50.1,ICD10 I50.9 |
| Heart Transplant | Heart Transplant | ICD10 Z94.1, ICD10 T86.2, Cptpb.33945, Echo Derived |
| HLHS | Hypoplastic Left Heart Syndrome | CCS 14.1.15, Echo Derived |
| HTN-PREG | Hypertension Complicating Pregnancy | CCS 11.3.3 |
| Hypertension | Hypertension | CCS 7.1 |
| Hypertensive Disease | Hypertensive Disease | ICD10 I10-I16 |
| Infectious Disease | Infectious Disease | ICD10 A00-B99 |
| Ischemic Heart Disease | Ischemic Heart Disease | ICD10 I20-I25 |
| Laterality Defects | Laterality Defects | Echo Derived |
| Milrinone | Milrionone | cui.52769 |
| Myocarditis | Myocarditis | CCS 7.2.2 |
| Nervous | Congenital Abnormalities of the Nervous System | CCS 14.4 |
| Norwood | Norwood Surgery | Echo Derived |
| Obesity | Obesity | CCS 3.11.2 |
| RV fxn decreased | Right Ventricle Function Decreased | Echo Derived |
| Skeletal | Congenital Abnormalities of the Skeletal System | CCS 14.5.17, CCS 14.5.15, CCS 14.5.13, CCS 14.5.14, CCS 14.5.9, CCS 14.5.18, CCS 14.5.16, CCS 14.5.24 |
| Skin | Congenital Abnormalities of Skin | CCS 14.5.23, CCS 14.5.22 |
| SND | Sinoatrial Node Dysfunction | CCS 7.2.9.6 |
| Tachycardia | Tachycardia | CCS 7.2.9.2 |
| Tacrolimus | Tacrolimus | cui.42316 |
| TGA | Transposition of the Great Arteries | CCS 14.1.1, Echo Derived |
| TOF | Tetralogy of Fallot | CCS 14.1.2, Echo Derived |
| Tricuspid Atresia | Tricuspid Valve Atresia | Echo Derived |
| TR | Tricuspid Regurgitation > moderate | Echo Derived |
| Viral Carditis | Viral Carditis | ICD10 B33.2 |
| VSD | Ventricular Septal Defect | CCS 14.1.3, Echo Derived |
